# Resting and functional pupil response metrics indicate features of reward sensitivity and Autism Spectrum Disorder in children

**DOI:** 10.1101/2020.01.24.20018648

**Authors:** Antoinette Sabatino DiCriscio, Vanessa Troiani

## Abstract

Altered motivational drives and aberrant reward system function may contribute to the social impairments observed in autism spectrum disorders (ASD). Pupil metrics have been highlighted as peripheral indicators of autonomic arousal and reward system function, specifically noradrenergic and dopaminergic activity that influence motivational drive states. However, research on individual differences in the neurobiological correlates of reward responsivity and clinically relevant features associated with ASD is sparse. The goal of the current study was to examine the relationship between measures of sensitivity to punishment and reward, ASD features, and resting as well as functional pupil response metrics across a clinically heterogeneous pediatric sample. We assessed whether quantitative features of reward sensitivity are linearly related to core clinical features of ASD. Pupil metrics were measured using a passive eye tracking task. Scores on a parent-report measure of punishment and reward sensitivity were found to be positively correlated with ASD features. Given these relationships, we assessed whether pupil measurements could be used as a neurobiological correlate of reward sensitivity and predictor of clinically significant ASD traits. In a logistic regression model, we find that the amplitude of pupil dilation, along with sex and full-scale IQ, could be used to correctly classify 84.9% of participants as having an ASD diagnosis versus not having an ASD diagnosis. This research highlights individual differences of reward sensitivity that scale with ASD features. Furthermore, reported results emphasize that functional pupil response metrics and other objective patient-level variables can be used together as predictors of ASD diagnostic status.

## BACKGROUND

It has been hypothesized that core diagnostic features of autism spectrum disorder (ASD) result from disruptions to neural networks associated with reward processing and motivation (Chevallier, Kohls, Troiani, Brodkin, & Schultz, 2012; Clements et al., 2018; Dichter, Richey, Rittenberg, Sabatino, & Bodfish, 2012; Kohls, Chevallier, Troiani, & Schultz, 2012; Mundy, Henderson, Inge, & Coman, 2007). This hypothesis has primarily been tested using functional magnetic resonance imaging (fMRI), in which participant’s brain response to various types of rewards are assessed (see Bottini, 2018; Clements et al., 2018 for review). The nature of the behavioral requirements to complete an fMRI experiment and the cognition required to assess participant’s understanding of abstract reward concepts (i.e. money) frequently limit participation in such studies to only high functioning children. In the current study, we assessed whether quantitative features of reward sensitivity are linearly related to core clinical features of ASD. First, we assessed reward sensitivity and ASD features in children with and without a diagnosis of ASD using behavioral measures (Sensitivity to Punishment and Sensitivity to Reward Questionnaire for Children, SPSRQ-C; and Social Responsiveness Scale, SRS). Based on previous research in this area (Luman, van Meel, Oosterlaan, & Geurts, 2012; L. Van den Berg et al., 2011), we predicted a significant linear relationship between SRS and SPSRQ-C scores. In Experiment 2, we explored the relationship between ‘resting’ or baseline pupil diameter (PD) and functional pupil response metrics collected from a passive eye tracking procedure, reward sensitivity, and ASD.

Pupil dynamics, including the pupillary light reflex (PLR), have been highlighted as a peripheral marker of underlying physiologic function as well as neurological and neuromuscular disease (Fotiou et al., 2007; Fotiou, Fountoulakis, Goulas, Alexopoulos, & Palikaras, 2000; Jain et al., 2011). The pupil has been interpreted as a peripheral neurobiological correlate of autonomic arousal and neurotransmitter release, specifically dopaminergic and noradrenergic activity (Bast, Poustka, & Freitag, 2018; Giza, Fotiou, Bostantjopoulou, Katsarou, & Karlovasitou, 2011; Hellmer & Nyström, 2017) that influence motivational drive states. Resting PD is thought to be a sensitive measure of synergistic sympathetic and parasympathetic activity within the ANS and related to tonic firing of neurons in the locus coeruleus (Anderson & Colombo, 2009; Bast et al., 2018; Fan, Miles, Takahashi, & Yao, 2009a; B.□; Laeng B.□;. Gredeback, G., 2012; Stern, Stern, Ray, & Quigley, 2001). Reactive changes in pupil diameter are influenced by numerous stimulus-driven, or external factors, including ambient light, motion, color and contrast (Ahern & Beatty, 1979; Birren, Casperson, & Botwinick, 1950; Ellis, 1981; Lobato-Rincón et al., 2014). However, changes in pupil response are also known to be influenced by arousal state, cognitive effort and visual attention (Binda, Pereverzeva, & Murray, 2014; Binda, Pereverzeva, & Murray, 2013; DiCriscio, Hu, & Troiani, 2018; Mather, Clewett, Sakaki, & Harley, 2015; Mathôt & Stigchel, 2015; Mathôt, van der Linden, Grainger, & Vitu, 2013). Thus, simple and reflexive changes in pupil diameter represent sensitive and reliable measures of cognitive processes as well as a proxy for underlying neurobiological function.

Outside of behavioral research studies on task- or stimulus-induced changes in pupil response in the context of reward (Bijleveld, Custers, & Aarts, 2009; S. Mardaga & Hansenne, 2009; Solange Mardaga & Hansenne, 2011; Matthys, Van Goozen, Snoek, & Van Engeland, 2004), baseline or resting PD has also been highlighted as a potential physiological marker for arousal state associated with cognitive processes including performance in reward-based paradigms (Aminihajibashi, Hagen, Foldal, Laeng, & Espeseth, 2019; Gilzenrat, Nieuwenhuis, Jepma, & Cohen, 2010; Unsworth & Robison, 2015).

Previous eye tracking and pupillometry research has reported differences in measures of resting PD in ASD (Anderson & Colombo, 2009; Anderson, Colombo, & Unruh, 2013; Blaser, Eglington, Carter, & Kaldy, 2014; Martineau et al., 2011), specifically reporting larger baseline pupil size in those individuals with ASD as compared to controls. These findings have been interpreted to reflect dysregulated arousal states within ASD that align with original hypotheses of autism as a disorder or atypical resting-state physiology (Hutt, Hutt, Lee, & Ounsted, 1964). Other research has focused specifically on the PLR, an automatic and reflexive pupil response following a brief flash of light that serves to control the amount of light falling on the retina. Within ASD, an atypical PLR has been reported (Dinalankara, Miles, Nicole Takahashi, & Yao, 2017; Fan et al., 2009a) and has been used to discriminate ASD patients from controls. In contrast to the PLR, recent work has focused on characterizing pupil adaptation metrics associated with ASD. Instead of a brief and transient stimulus, sustained monochromatic stimuli are presented and functional response metrics including the amplitude peak pupil response are extracted. DiCriscio and Troiani (2017), using a simple eye tracking paradigm, assessed characteristic patterns of pupil dilation and constriction in response to alternating dark and light stimuli across a clinically heterogeneous cohort of children (i.e. with and without ASD and with a broad range of cognitive abilities). Individual differences in functional pupil response metrics were found to be associated with a quantitative measure of ASD traits. Taken together, this research highlights pupillary metrics as potential neurobiologic correlates of core clinical features of ASD. However, it remains unclear how resting PD and functional pupil responses are associated with other cooccurring cognitive features, such as reward sensitivity, that may be associated with core clinical features of ASD.

The measure we employ here to assess sensitivity to reward and punishment is based on Gray’s Reinforcement Sensitivity Theory (RST), which conceptualizes individual differences in variation in the brain’s sensitivity to punishing and reinforcing stimuli (Gray, 1976, 1982). Revised versions of Gray’s original RST (Cogswell, Alloy, van Dulmen, & Fresco, 2006; Cooper, Smillie, & Jackson, 2008; Gray & McNaughton, 2000; McNaughton & Corr, 2004; Smillie, Pickering, & Jackson, 2006) have led to the current understanding of the behavioral inhibition (BIS) and behavioral approach systems (BAS) as interdependent neurobiological systems that influence human behavior in response to signals of reward and punishment.

When considering the RST framework and disruptions to reward and punishment sensitivity in ASD, one could hypothesize a link between disturbances in appetitive behaviors associated with the BAS and symptoms related to social approach. However, research in joint attention across ASD suggests a more complex relationship between BIS/BAS function, approach/avoidance behaviors and a broad range of symptoms associated with social impairment. Reduction in the *initiation* of joint attention in ASD (Dawson et al., 2005; Kasari, Sigman, Mundy, & Yirmiya, 1990; Meindl & Cannella-Malone, 2011; P. C. Mundy et al., 2007; P. Mundy, Sigman, & Kasari, 1992; Peter Mundy, 1995), suggests that disturbances in BAS activity and atypical social approach behaviors contribute to joint attention deficits. Deficits in *responding* to joint attention bids from others have also been described (Loveland & Landry, 1986; Whalen & Schreibman, 2003) and align with disruptions in BIS activity and atypical avoidance behaviors that may contribute to diminished social orienting, symptoms of social withdrawal, aloof behaviors, (Chevallier et al., 2012; Gadow & Garman, 2018; Mikami, Miller, & Lerner, 2019) and comorbid symptoms of anxiety in ASD (Leyfer et al., 2006). Thus, it is unclear whether social features central to ASD are driven solely by BIS or BAS dysfunction. Furthermore, the restricted and repetitive behavior symptom domain of ASD cannot be fully accounted for within the framework of disturbed approach behaviors and instead may suggest that such symptoms emerge from atypical sensitivity to signals of punishment that call for behavioral modification (Geurts, Corbett, & Solomon, 2009; Yerys et al., 2009). Thus, the heterogeneous and variable phenotypic expression of multiple ASD symptom domains may be attributed to variability in both approach and avoidance behaviors across individuals.

The RST framework has been previously related to peripheral somatic markers of autonomic response (Colder et al., 2011; De Pascalis, Fiore, & Sparita, 1996; S. Mardaga & Hansenne, 2009, 2009; Solange Mardaga & Hansenne, 2011; Norris, Larsen, & Cacioppo, 2007); however, has not been related to pupillary measurements. Pupil size is influenced by a variety of factors including arousal, visual attention, learning and reward responses (Ahern & Beatty, 1979; P. Binda et al., 2014; Paola Binda & Murray, 2015; Paola Binda et al., 2013; Kahneman & Beatty, 1966; B. Laeng, Ørbo, Holmlund, & Miozzo, 2011; Mathôt, Dalmaijer, Grainger, & Van der Stigchel, 2014; Mathôt et al., 2013) and is linked to activity in a network of brain regions and brainstem nuclei (Beatty & Lucero-Wagoner, 2000; Corbett, Mendoza, Abdullah, Wegelin, & Levine, 2006; Corbett, Mendoza, Wegelin, Carmean, & Levine, 2008; Hou, Langley, Szabadi, & Bradshaw, 2007; Stenberg, 2007). Previous work showing greater baseline PD (Anderson & Colombo, 2009; Anderson et al., 2013) and meaningful variability in reflexive pupil response (DiCriscio & Troiani, 2017; Fan et al., 2009a) in ASD has suggested atypical or discordant autonomic arousal may drive these differences; however, it remains unclear how pupil measurements may be associated with individual differences in RST activity.

We aimed to assess the relationship between sensitivity to punishment and reward and ASD features in a clinically heterogeneous pediatric sample of children with and without ASD, including children with mild to moderate intellectual disability (Russell et al., 2019). The present study assesses resting PD and functional pupil response metrics in the absence of specific stimuli and/or subjective content. Based on previous work in ASD and other neurodevelopmental phenotypes (Anderson & Colombo, 2009; DiCriscio & Troiani, 2017; Nuske, Vivanti, & Dissanayake, 2014b), we predicted significant relationships between pupil measurements, SPSRQ-C and SRS scores. Specifically, we predicted that resting PD and the amplitude of pupil response would scale with the presence of ASD features. We then assessed whether measures of PD could be used as neurobiological correlates of reward sensitivity and a predictor of ASD diagnostic status.

## EXPERIMENT 1

### STUDY 1

#### METHODS

##### Participants and General Procedure

Participants (N=89; mean age= 8.48 ± 2.25; 54 males) included children 5 thru 14 years of age. We used a broad recruitment strategy in order to obtain a cohort with a wide range of autism traits. This included identifying participants based on patient referral to our neurodevelopmental pediatric clinic, as well as from health system wide advertisement. Participants were recruited from our clinic via enrollment in our clinic’s research protocol, which enables access to electronic health record variables and re-contact for additional research. Of the entire sample described above, n=43 individuals (48% of sample) had a clinical diagnosis of ASD (34 males). ASD diagnoses were determined based on a team of clinicians and support staff at a neurodevelopmental pediatric clinic that specializes in ASD and neurodevelopmental disorders. Participants were recruited as a part of a larger study that also gathered eye tracking (see Experiment 2) and additional cognitive and behavioral data. On the day of research testing, all participants completed a cognitive assessment to document IQ (WASI-II: Wechsler abbreviated scale of intelligence, 2^nd^ edition; (Wechsler & Hsiao-pin, 2011) (K-BIT: Kaufman Brief Intelligence Test, 2^nd^ edition; (Kaufman & Kaufman, 2004). If an IQ test was completed during their clinic appointment that day (n=21), we used the clinically ascertained IQ score. All participants assented to protocols approved by the institutional review board (IRB) at the authors’ home institution.

##### Parent-report Measures and Scoring

Parents were asked to complete parent-report forms of questionnaires (e.g. reporting based on their child’s behavior). Questionnaires were administered electronically via laptop computer supplied to parents at the research visit. Scores on all parent-report and cognitive measures can be found in Table 1.

**Table 1.**
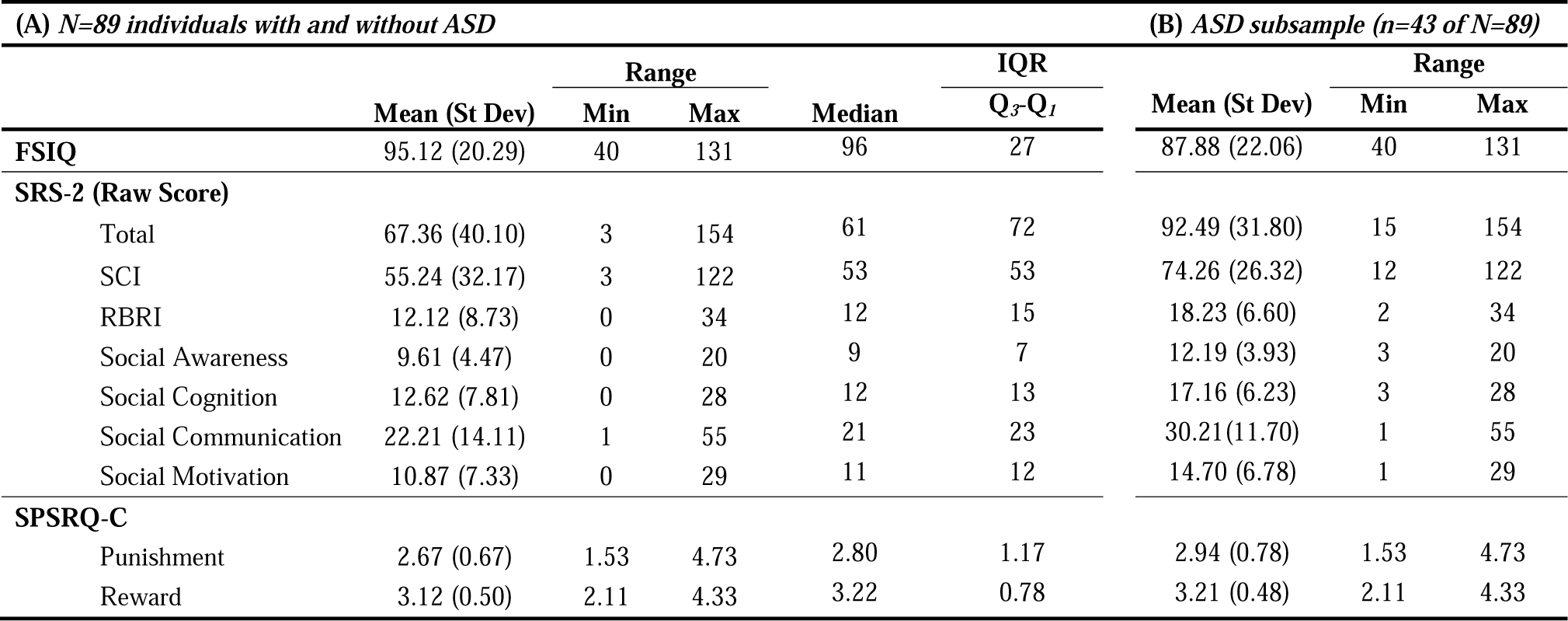
(A) Range and means (SDs) for cognitive and parent-report measures (N=89) as well as (B) Range and means (SDs) for subset of individuals with ASD Dx (i.e. n=43 of N=89).

###### Social Responsiveness Scale-2^nd^ Edition (SRS-2)

The Social Responsiveness Scale-2^nd^ Editions (SRS-2; Constantino et al., 2003; Frazier et al., 2013) is most frequently used as a parent-report measure assessing the presence and severity of symptoms of social impairment associated with ASD. In addition to a Total score reflecting overall impairments and social communication impairments (SCI), the SRS-2 generates scores across five subscales (Social Cognition, Social Motivation, Social Awareness, Social Communication, and Restricted Interests and Repetitive Behaviors).

###### Sensitivity to Punishment and Sensitivity to Reward Questionnaire/- for Children (SPSRQ-C)

The Sensitivity to Punishment and Sensitivity to Reward Questionnaire for Children (SPSRQ-C; (Colder & O’Connor, 2004) is a 33-item parent-report measure with responses provided on a 5-point Likert scale, ranging from 1 (strongly disagree) to 5 (strongly agree). A 2-factor model of the SPSRQ-C results in two subscales: Punishment and Reward (Colder & O’Connor, 2004; Luman et al., 2012). The Punishment subscale is theoretically linked to the BIS and the Reward subscale is linked to the BAS. Group average SPSRQ-C scores can be found in Table 1. Internal consistency of the SPSRQ-C, based on Cronbach’s alpha, within our sample is reported in Table 2 and is consistent with previous research (Ezpeleta, Granero, de la Osa, & Domènech, 2017; I. Van den Berg, Franken, & Muris, 2010). Additionally, we assessed the internal consistency of the SPSRQ-C separately for our ASD and non-ASD subsamples and found these reliability measures to be consistent across both groups.

**Table 2.** Internal Consistency (Cronbach’s alpha, α) of SPSRQ-C Punishment and Reward (Entire sample, N=89; ASD subsample, n=43; Non-ASD subsample, n=46)

| | Cronbach's $\alpha$ | | |
| --- | --- | --- | --- |
|  | Entire Sample | Sample separated by Dx |  |
|  |  | ASD<br>*n*=43 | Non-ASD<br>*n*=46 |
| **SPSQRQ-C** | *N*=89 |  |  |
| **Punishment** | 0.886 | 0.878 | 0.888 |
| **Reward** | 0.789 | 0.808 | 0.786 |

##### Data Analysis

Our main analyses focused on (1) investigating the relationship between individual differences in autism features and measures of reward sensitivity and (2) identifying whether measures of reward sensitivity significantly predicted the presence of an ASD diagnosis. A partial correlation, controlling for age, FSIQ, and sex, between SRS-2 and SPSRQ-C scores was used to assess the relationship between reward sensitivity and autism features. SPSRQ-C scores, as well as FSIQ and sex, were entered into a binary logistic regression to identify whether behavioral features of reward and punishment sensitivity significantly predicted diagnosis. It is important to note that raw SRS-2 scores were used for all primary analyses in order to provide greater variability of scores at the lower and higher end of the measure across our clinically heterogeneous sample (Bölte, Poustka, & Constantino, 2008; Constantino & Gruber, 2005; Duvekot, van der Ende, Verhulst, & Greaves-Lord, 2015; Moreno-De-Luca et al., 2015).

Before completing our primary analyses, we determined whether there were relationships that may impact interpretation between our demographic variables (i.e. child’s age and FSIQ) and parent-report measures. Age was not found to be related to SRS scores (*p*’s>0.40) nor SPSRQ-C scores (*p*’s>0.88). FSIQ was found to be related to SRS Total score (r= −0.239, *p*=0.024) as well as all subscale scores (*p*’s<0.04) with the exception of the Social Awareness subscale (r= −0.18, *p*= 0.09, NS) and the Social Motivation subscale (r= −0.20, *p*=0.06, NS). FSIQ was not found to be related to SPSRQ-C scores (*p*’s>0.26).

We also assessed the distribution of parental report measures. SRS Total raw scores deviated from a normal distribution based on results of Shapiro-Wilks tests of normality (*p*=0.001). SPSRQ-C Reward (*p*= 0.207, NS) and SPSRQ-C Punishment (*p*=0.09) did not deviate from a normal distribution1. SRS Total raw scores for the entire sample demonstrated a bimodal distribution. We also assessed the distribution of SRS raw scores separately for those with and without an ASD diagnosis. SRS raw scores in those individuals with an ASD diagnosis demonstrated a moderately left skewed distribution, −0.678 (SE= 0.36), while those without an ASD diagnosis demonstrated a moderately right skewed distribution, 0.810 (SE=0.35). Follow-up t-tests comparing SRS scores between individuals with and without an ASD diagnosis indicated significantly higher SRS Total raw scores in those individuals with an ASD diagnosis (mean=92.49 ± 31.80) than those without (mean=41.61 ± 28.82), t(87)= −7.918, *p*<0.0001. Scores on all parent-report measures (i.e. SRS and SPSRQ-C scores) were then transformed into z-scores and standardized scores were used across all primary analyses below.

## RESULTS

### Relationship between reward responsivity and autism features

Given the distribution of SRS scores across our sample and the violation of normality, we examined the relationship between SRS scores and SPSRQ-C scores via nonparametric Spearman’s, rank correlation. SRS Total raw score was found to be significantly related to SPSRQ-C Punishment (r= 0.401, *p*<0.0001) as well as SPSRQ-C Reward (r=0.299, *p*=0.004) (see Figure 1). SPSRQ-C Punishment and SPSRQ-C Reward were also found to be significantly related to all SRS subscales (*p*’s<0.016, see Table 3).

**Table 3.**
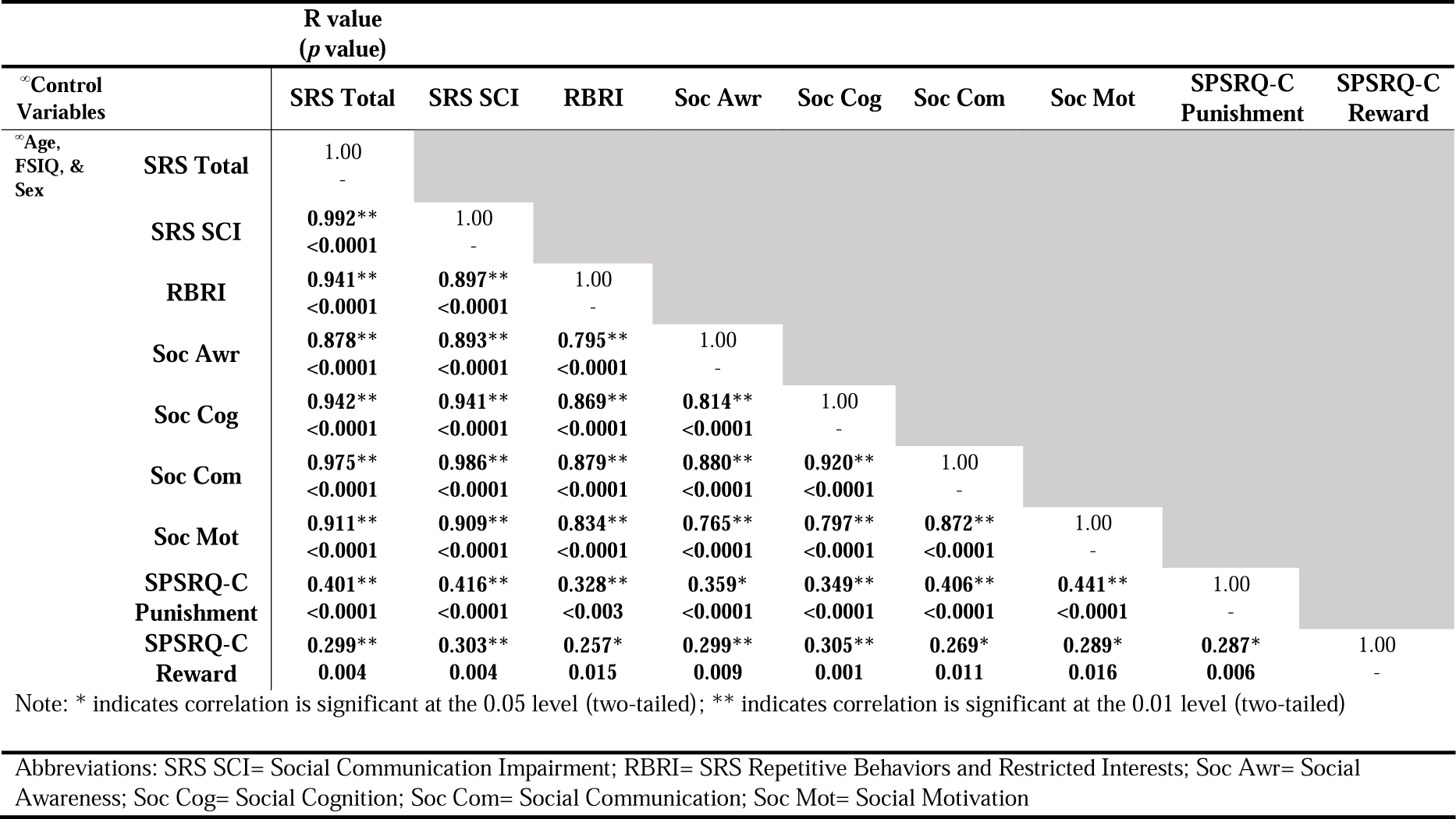
Spearman’s, rank correlation between SRS raw scores and SPSRQ-C Punishment and Reward subscale scores (based on z-score transformations on all parent-report measures).

**Figure 1.**
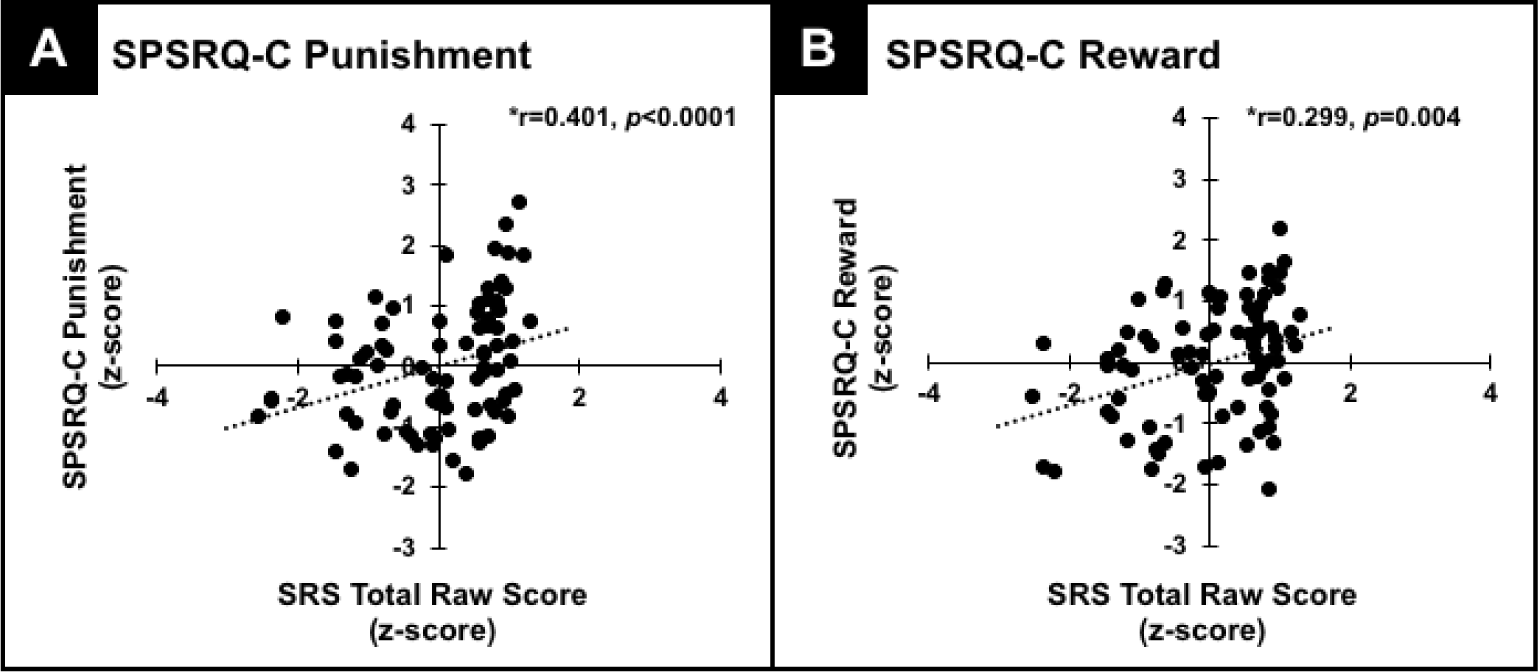
Results of a nonparametric, Spearman’s rank correlation (age, FSIQ, and sex) indicating a significant relationship between (A) SPSRQ-C Punishment (z-score) and SRS Total raw score (z-score) (r=0.401, *p*<0.0001); (B) SPSRQ-C Reward (z-score) and SRS Total raw score (z-score) (r=0.299, *p*=0.004).

### Binary Logistic Regression

A binary logistic regression was performed to assess the effects of SPSRQ-C Punishment, SPSRQ-C Reward, FSIQ, age, and sex on predicting the presence of an ASD diagnosis. The logistic regression model was significant (χ2(5)= 29.758, *p*<0.0001, Nagelkerke R^2^= 0.38) and correctly classified 69.7% of cases as having an ASD diagnosis. FSIQ, sex, and SPSRQ-C Punishment were noted to be significant predictors of diagnosis in our sample (see Table 4 for complete results).

**Table 4.**
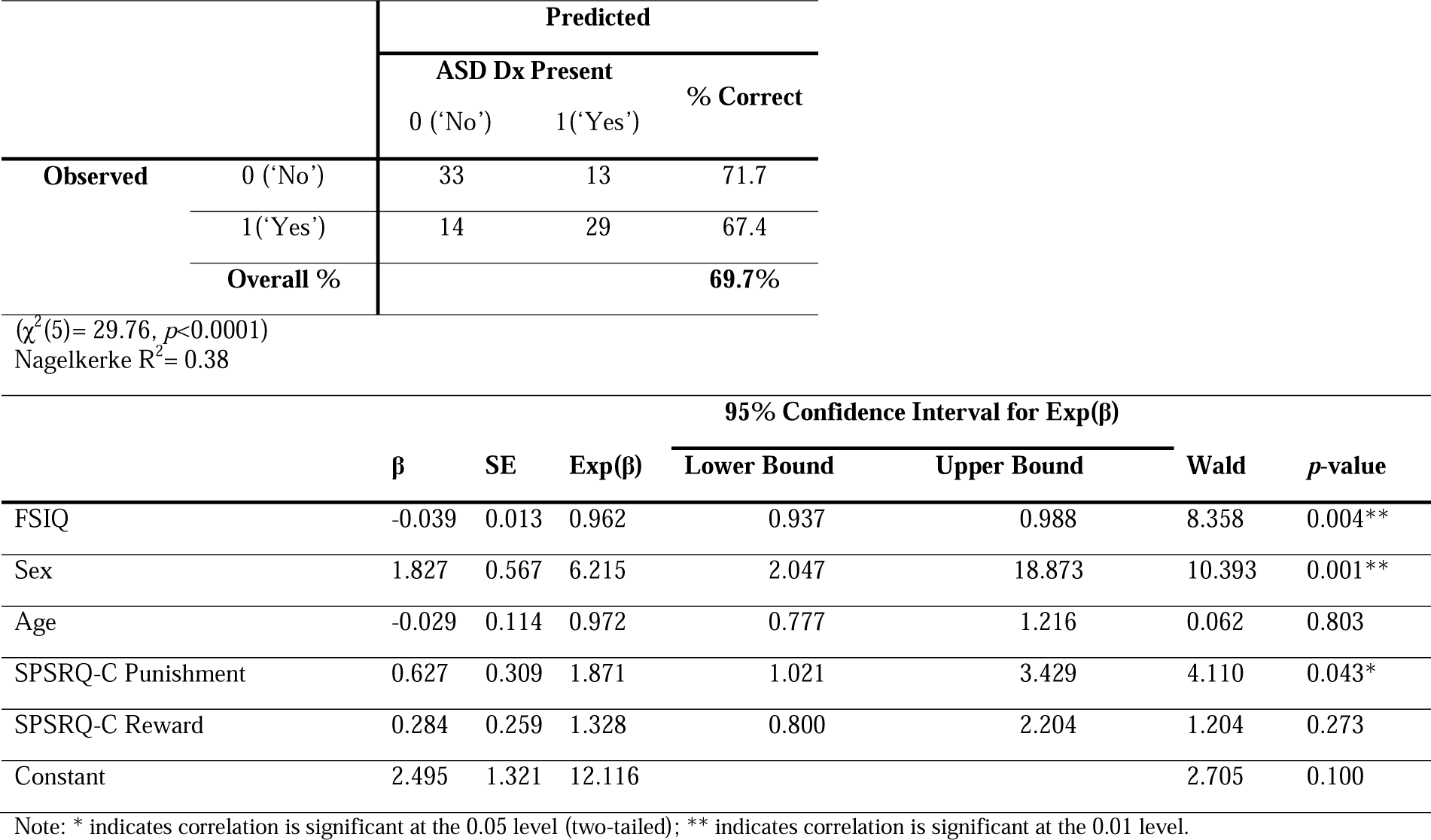
Binary Logistic Regression predicting ASD Dx including FSIQ, SPSRQ-C Punishment, and Reward subscales, and Sex as predictors.

### Experiment 1 Conclusions

Results outlined above demonstrate a linear relationship between individual differences in reward and punishment sensitivity based on parental report questionnaires and ASD. While this research contributes to the growing body of literature on atypical reward sensitivity and core ASD features, the results described above are based on parent-report measures. Outside of the use of parental-report metrics, neuroimaging methods have consistently documented atypical reward response in ASD; however, neuroimaging is expensive and difficult to implement in individuals with significant behavioral and cognitive impairments. Thus, there is a need for sensitive and objective measures of reward sensitivity associated with clinically relevant ASD features. In the next experiment, we extended previous research on differences in baseline PD in ASD (Anderson & Colombo, 2009; Fan, Miles, Takahashi, & Yao, 2009b) and meaningful variability in functional pupil response metrics (i.e. characteristic patterns of reflexive dilation and constriction) associated with clinical ASD features (DiCriscio & Troiani, 2017). We explored whether resting PD and functional pupil changes in response to alternating light and dark conditions were associated with reward and punishment sensitivity and quantitative measures of ASD features.

## EXPERIMENT 2

### METHODS

#### Participants and General Procedure

In addition to parent-report and cognitive measures outlined in Experiment 1 (i.e. SPSRQ-C and SRS), a subset of participants (N=73, mean age = 8.68 ± 2.186, n= 45 males) successfully completed an eye tracking session at the time of their in-person research appointment (i.e. n=4 participants from our sample in Experiment 1 did not complete the eye tracking task, n=12 participants from Experiment 1 were excluded from the analysis for Experiment 2 due to unsuccessful completion of our eye tracking task)^2^. Of the larger sample included in Experiment 2, n=43 individuals (58% of sample) had a diagnosis of ASD (34 males). See Table 5 for demographics and scores on parent-report measures for larger sample and ASD subsample included in Experiment 2.

**Table 5.** (A) Range and means (SDs) for subset with parent-report measures *and* eye tracking data that were included in analysis for Experiment 2 (N=73) as well as (B) Range and means (SDs) for subset of individuals with ASD Dx (i.e. n=43 of N=73).

| (A) N=73 individuals with and without ASD |  |  |  |  |  | (B) ASD subsample (n=43 of N=73) |  |  |
| --- | --- | --- | --- | --- | --- | --- | --- | --- |
|  | Mean (St Dev) | Range |  | Median | IQR<br>Q<sub>3</sub>-Q<sub>1</sub> | Mean (St Dev) | Range |  |
|  |  | Min | Max |  |  |  | Min | Max |
| ***IQ & Parent-Report Measures*** |  |  |  |  |  |  |  |  |
| **FSIQ** | 95.53 (20.60) | 40 | 131 | 100 | 29 | 87.88 (22.06) | 40 | 131 |
| **SRS-2 (Raw Score)** |  |  |  |  |  |  |  |  |
| Total | 73.31 (39.08) | 6 | 154 | 82 | 65 | 92.49 (31.80) | 15 | 154 |
| SCI | 59.90 (31.31) | 5 | 122 | 67 | 50 | 74.26 (26.32) | 12 | 122 |
| RBRI | 13.41 (8.68) | 0 | 34 | 15 | 15 | 18.23 (6.60) | 2 | 34 |
| Social Awareness | 10.35 (4.29) | 0 | 20 | 10 | 7 | 12.19 (3.93) | 3 | 20 |
| Social Cognition | 13.72 (7.67) | 0 | 28 | 14 | 13 | 17.16 (6.23) | 3 | 28 |
| Social Communication | 24.08 (13.90) | 1 | 55 | 26 | 22 | 30.21(11.70) | 1 | 55 |
| Social Motivation | 11.75 (7.38) | 1 | 29 | 12 | 13 | 14.70 (6.78) | 1 | 29 |
| **SPSRQ-C** |  |  |  |  |  |  |  |  |
| Punishment | 2.75 (0.76) | 1.53 | 4.73 | 3.22 | 1.10 | 2.94 (0.78) | 1.53 | 4.73 |
| Reward | 3.16 (0.48) | 2.11 | 4.33 | 2.73 | 0.64 | 3.21 (0.48) | 2.11 | 4.33 |
| ***Pupil Measures*** |  |  |  |  |  |  |  |  |
| Resting PD (mm) | 3.86 (0.67) | 2.72 | 5.83 | 3.73 | 0.91 | 4.02 (0.72) | 2.77 | 5.83 |
| Amplitude of Dilation- $A_D$ (mm) | 1.34 (0.64) | 0.16 | 2.44 | 1.42 | 1.16 | 1.01 (0.08) | 0.16 | 2.07 |
| Amplitude of Constriction- $A_C$ (mm) | 1.36 (0.85) | 0.12 | 2.92 | 1.27 | 1.65 | 0.94 (0.11) | 0.12 | 2.76 |

Prior to the start of the eye tracking task, a gray screen was presented for 10 sec from which resting PD was extracted. After this gray screen, alternating dark (i.e. black screen) and light (i.e. white screen) stimuli were displayed across a ∼2.5-minute task (24 trials total; 12 each for dark and light conditions). Each stimulus screen was presented for 5 seconds. Participants were instructed to remain still and to maintain gaze in the center location of the screen. See (DiCriscio & Troiani, 2017) for additional details regarding the eye tracking paradigm. See Table 4 for descriptive statistics for average resting PD and amplitude of pupil responses for larger sample and ASD subsample included in Experiment 2. See Table 4 for descriptive statistics for average resting and functional measures of pupil response for larger sample and ASD subsample included in Experiment 2. All analyses outlined below were repeated, including the average percent of trials successfully compelted as a covariate, and yielded similar results.

#### Eye tracking-resting PD and pupil response metrics

Data was exported from Tobii Studio and analysis proceeded using MATLAB and SPSS. In the event of missing data from one pupil, missing values were replaced with the recorded value for the other eye. In the event of missing values for both eyes, a linear interpolation was used. Pupil data was averaged and smoothed using a low pass (15 Hz) filter. Average resting PD was extracted from the 10 second gray screen presented at the start of the task.

Differences in sustained pupil response across dark and light conditions (i.e. the *amplitude* of dilation-*A*_D_ or constriction-*A*_C_) were measured as changes in pupil diameter relative to the average baseline pupil diameter (resting PD) measured at the start of the task. Measures of reflexive changes in pupil response transitioning from one condition to the other were then averaged across each condition. See Figure 1 (a) dark condition and (b) light condition. DiCriscio and Troiani (2017) extracted amplitude metrics *as well as* latency to reach maximum dilation and constriction; however, results indicated amplitude metrics to be a significant predictor of ASD features as compared to latency metrics. Thus, we focused analysis on pupil amplitude measures in the current study.

#### Data Analysis

We next focused on (1) investigating the relationship between individual differences in resting PD, functional pupil response metrics (*A*_*C*_, *A*_*D*_), autism features and measures of reward sensitivity, and (2) identifying whether resting and/or functional pupil metrics significantly predicted the presence of an ASD diagnosis. Pupil metrics, as well as FSIQ and sex, were entered into a binary logistic regression to identify whether these objective metrics significantly predicted the presence of an ASD diagnosis. Pupil metrics was transformed into z-scores and standardized measures were used across all analyses reported below. Please note that all pupil metrics (resting PD, *A*_*C*_, and *A*_*D*_) deviated from a normal distribution based on results of Shapiro-Wilks tests of normality (*p’s*<0.005).

## RESULTS

### Relationship between resting PD, A_C_, A_D_, reward responsivity and SRS scores in children

Resting PD was found to be significantly correlated with SRS Total raw score (r=0.282, *p*=0.014) and SPSRQ-C Reward (r=0.360, *p*=0.002) (see Figure 2A). A relationship between resting PD and SPSRQ-C Punishment approached but did not reach significance (r=0.20, *p*>.08, NS). Complete results from this correlation are reported in Table 6. *A*_*D*_ was found to be related to SRS Total raw score (r= −0.537, *p*<0.0001), SPSRQ-C Punishment (r= −0.393, *p*=0.001), as well as SPSRQ-C Reward (r= −0.237, *p*=0.049) (see Figure 2B). Similarly, *A*_*C*_ was also found to be significantly related to SRS Total raw score (r= −0.515, *p*<0.0001), SPSRQ-C Punishment (r= −0.495, *p*<0.0001), and SPSRQ-C Reward (r= −0.309, *p*=0.010) (see Figure 2C).

**Table 6.**
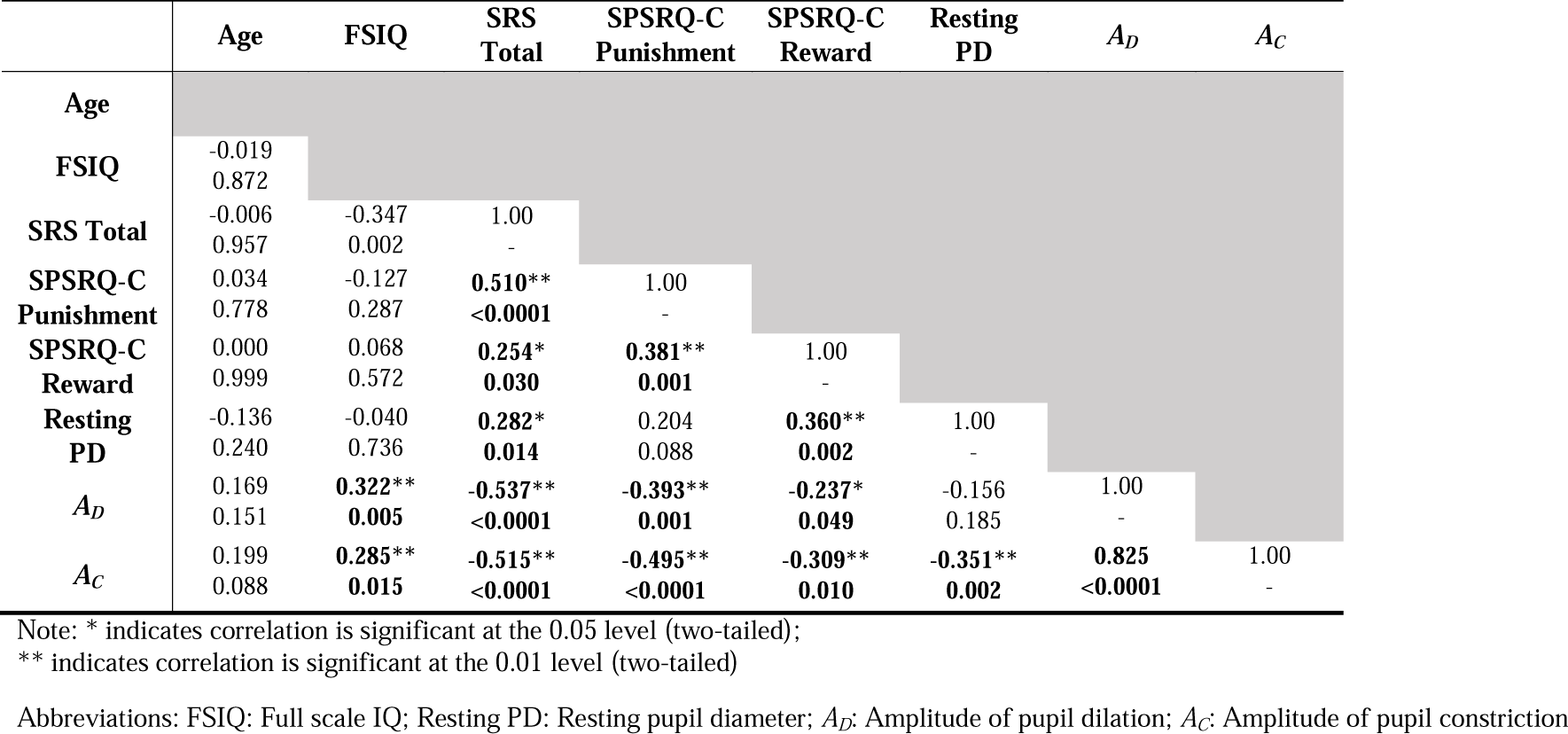
Spearman, rank correlation between Age, FSIQ, SRS raw scores, SPSRQ-C Punishment and Reward subscale scores, and metrics of PD.

| | Age | FSIQ | SRS<br>Total | SPSRQ-C<br>Punishment | SPSRQ-C<br>Reward | Resting<br>PD | $A_D$ | $A_C$ |
| --- | --- | --- | --- | --- | --- | --- | --- | --- |
| **Age** |  |  |  |  |  |  |  |  |
| **FSIQ** | -0.019<br>0.872 |  |  |  |  |  |  |  |
| **SRS Total** | -0.006<br>0.957 | -0.347<br>0.002 | 1.00<br>- |  |  |  |  |  |
| **SPSRQ-C**<br>**Punishment** | 0.034<br>0.778 | -0.127<br>0.287 | **0.510\*\***<br>**<0.0001** | 1.00<br>- |  |  |  |  |
| **SPSRQ-C**<br>**Reward** | 0.000<br>0.999 | 0.068<br>0.572 | **0.254\***<br>**0.030** | **0.381\*\***<br>**0.001** | 1.00<br>- |  |  |  |
| **Resting**<br>**PD** | -0.136<br>0.240 | -0.040<br>0.736 | **0.282\***<br>**0.014** | 0.204<br>0.088 | **0.360\*\***<br>**0.002** | 1.00<br>- |  |  |
| $A_D$ | 0.169<br>0.151 | **0.322\*\***<br>**0.005** | **-0.537\*\***<br>**<0.0001** | **-0.393\*\***<br>**0.001** | **-0.237\***<br>**0.049** | -0.156<br>0.185 | 1.00<br>- | |
| $A_C$ | 0.199<br>0.088 | **0.285\*\***<br>**0.015** | **-0.515\*\***<br>**<0.0001** | **-0.495\*\***<br>**<0.0001** | **-0.309\*\***<br>**0.010** | **-0.351\*\***<br>**0.002** | **0.825**<br>**<0.0001** | 1.00<br>- |

Note: \* indicates correlation is significant at the 0.05 level (two-tailed);
\*\* indicates correlation is significant at the 0.01 level (two-tailed)
Abbreviations: FSIQ: Full scale IQ; Resting PD: Resting pupil diameter; $A_D$ : Amplitude of pupil dilation; $A_C$ : Amplitude of pupil constriction

**Figure 2.**
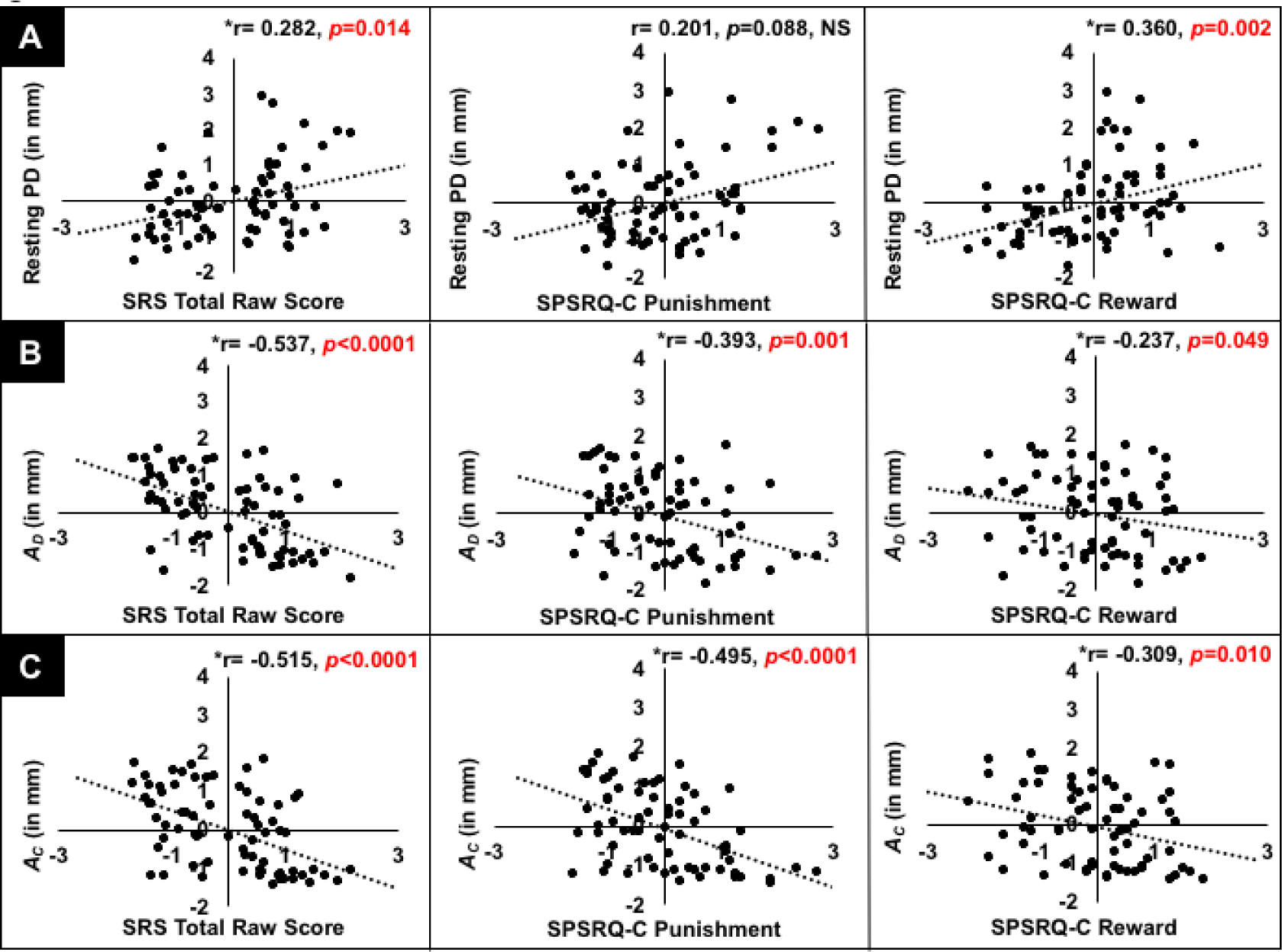
Results of a nonparametric, Spearman’s rank correlation (age, FSIQ, and sex) indicating a significant relationship between (A) Resting PD (Ø), SPSRQ-C Punishment, SPSRQ-C Reward, and SRS Total raw score (z-scores); (B) Amplitude of dilation during dark adaptation (*A*_*D*_) SPSRQ-C Punishment, SPSRQ-C Reward, and SRS Total raw score (z-scores); and (C) Amplitude of constriction during light adaptation (*A*_*C*_) SPSRQ-C Punishment, SPSRQ-C Reward, and SRS Total raw score (z-scores).

These results align with the correlations reported in Experiment 1 and suggest a novel relationship between resting PD, function pupil response metrics, quantitative measures of reward sensitivity, and ASD features. Complete results from a Spearman’s rank, nonparametric correlation, including SRS subscale scores, are reported in correlation tables provided as supplementary material for the larger sample (N=73) (see Supplementary Table 1) as well as our ASD subsample (e.g. n=43 of 73) (see Supplementary Table 2).

### Binary Logistic Regression

Given the results reported above, we explored whether resting and functional pupil response metrics could be substituted into regression analyses and function as neurophysiological correlates of SPSRQ-C scores that could predict ASD diagnostic status. A binary logistic regression was performed to assess the effects of resting PD, *A*_*D*_, *A*_*C*_, FSIQ, age and sex on predicting the presence of an ASD diagnosis. The logistic regression model was significant (χ2(6)= 51.80, *p*<0.0001, Nagelkerke R^2^= 0.68) and correctly classified 84.9% of cases as having an ASD diagnosis. *A*_*D*_, sex, and FSIQ were associated with the presence of clinically significant ASD symptoms in children (see Table 7).

**Table 7.**
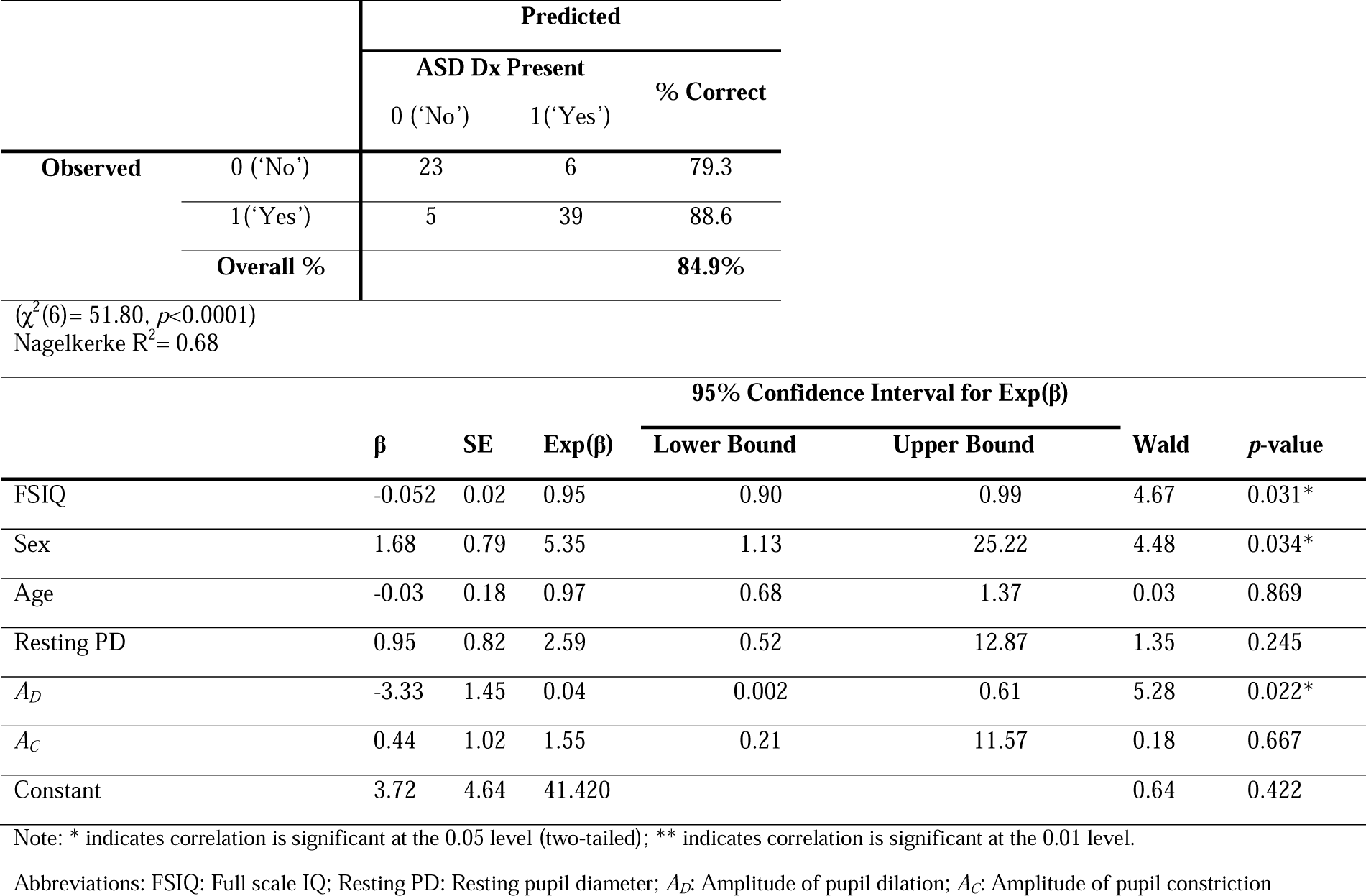
Binary Logistic Regression predicting ASD Dx including FSIQ, Sex, Age, Resting PD, *A*_*D*_, and *A*_*C*_ as predictors.

A model including resting and functional metrics of pupil response indicated that measures of pupil dilation during dark adaptation, as compared to resting PD and pupil response during light adaptation, are a significant predictor of diagnosis. We wish to highlight that our measure of resting PD in the current study parallels measures of baseline PD described in previous pupil research. As compared to measures of functional pupil changes assessed in response to alternating stimulus displays, resting PD or baseline PD is extracted in the absence of a stimulus, task, or visual input. Previous eye tracking and pupillometry studies in ASD have noted differences in baseline PD (Anderson & Colombo, 2009; Anderson et al., 2013; Blaser et al., 2014; Martineau et al., 2011; Nuske, Vivanti, & Dissanayake, 2014a) and broadly interpreted to reflect ANS activity and arousal (Anderson & Colombo, 2009; Nuske et al., 2014a) Given this previous research, we also ran supplementary analyses to explore whether resting PD, along with patient level variables, could be used a significant predictor of ASD diagnostic status. Results from a binary logistic regression including resting PD, age, sex, and FSIQ are reported in supplementary material.

## DISCUSSION

The current research aimed to characterize the linear relationship between measures of reward and punishment sensitivity, eye tracking metrics including resting PD and functional pupil response, and quantitative measures of ASD features. Specifically, we aimed to explore whether PD could function as peripheral objective measures of reward sensitivity that differentiates individuals with and without a diagnosis of ASD. We report significant relationships across SPSRQ-C and SRS scores as well as pupil measurements in a heterogeneous sample of children with and without ASD, demonstrating that individual differences in measures of punishment and reward sensitivity scale with the presence of ASD features.

We report a significant linear relationship between measures of punishment and reward sensitivity and SRS scores. Specifically, results from a binary logistic regression highlighted SPSRQ-C Punishment as a significant predictor of an ASD diagnosis. The Punishment subscale of both the SPSRQ and SPSRQ-C is theoretically linked to the BIS, reflecting behavioral inhibition, avoidance, and sensitivity to signals of punishment, non-reward, denial, and novelty (Luman et al., 2012; Vandeweghe, Vervoort, Verbeken, Moens, & Braet, 2016). Increased sensitivity to signals of negative reinforcement would result in altered behavioral inhibition and/or negative affective or behavioral responses (i.e. withdraw, fear or anxiety) to novel situations and social interactions (Carver & White, 1994; Jeffrey A. Gray, 1994). This definition aligns with core diagnostic features associated with ASD, including diminished social reciprocity, social orienting, reduced response to bids for joint attention, and observations of co-occurring anxiety, social withdraw and aloof behavior (Chevallier et al., 2012; Constantino, 2011; Mundy et al., 2007; Sherer & Schreibman, 2005; South, Dana, White, & Crowley, 2011).

Thus, behavioral features associated with behavioral inhibition and punishment sensitivity may significantly influence the development and clinical presentation of core autism features.

Of particular interest in the current study was the linear relationship between measures of pupil adaptation (amplitude of dilation), reward and punishment sensitivity, and ASD features. Given the significant relationships between SRS and SPSRQ-C scores and PD, resting PD and functional pupil metrics were substituted into a binary logistic regression for the parental report measures. In this adapted regression model, the amplitude of pupil dilation during dark adaptation, FSIQ, and sex could be used to predict ASD diagnostic status. These results emphasize that individual differences in reward sensitivity may be tied to physiologic indicators (pupil metrics) that can be objectively measured and potentially predictive of diagnosis. Thus, eye tracking technology and passive paradigms with minimal task demands may be used to extract meaningful oculomotor metrics that may serve as useful proxies for neurobiological correlates of co-occuring cognitive features such as reward sensitivity and autism features in children with ASD.

Differences in pupil dynamics have been noted in ASD. Atypical baseline PD, prior to the onset of a stimulus, have been reported in individuals with ASD as compared to controls and shown to be a significant predictor of diagnostic group membership (Anderson & Colombo, 2009; Lynch, James, & VanDam, 2018). Specifically, individuals with ASD exhibit larger resting PD as compared to peers (Anderson & Colombo, 2009; Blaser et al., 2014). Other studies have demonstrated differences in pupil response metrics during cognitive and perceptual tasks (Blaser et al., 2014; Martineau et al., 2011; Nuske, Vivanti, & Dissanayake, 2016). However, a majority of this previous work focused on comparisons between ASD and matched control groups and did not explore the relationship between pupil measures and individual differences in ASD features and co-occurring traits (DiCriscio, Hu, & Troiani, 2019b, 2019a). The work presented here extends previous research (DiCriscio & Troiani, 2017) assessing meaningful variability in pupil adaptation metrics associated with quantitiative measures of clinically relevant ASD traits. Specifically, the current research demonstrates the utility of functional pupil response metrics and patient level variables as potential diagnostic predictors. Additionally, to our knowledge, this is the first study to characterize the link between resting and functional pupil response metrics and quantitative features of reward sensitivity in ASD.

We wish to highlight that Reward and Punishment subscales were significantly correlated across Experiments 1 and 2 which is consistent with previous research that has reported significant relationships between the subscales of the SPSRQ-C (Van den Berg et al., 2010) as well as relationship between the BIS/BAS. More modern interpretations of Gray’s original RST framework hypothesizes interdependent biological subsystems (Corr, 2002) that modulate responses to signals of reward and punishment. Research in the general population suggests that there is meaningful variability in the function and sensitivity of these behavioral response systems (Ezpeleta et al., 2017). Thus, there may exist a broad spectrum of BIS/BAS response profiles (i.e. low-BIS/high-BAS to high-BIS/low-BAS) rather than mututally exclusive behavioral components that can be singularly assessed. Additional research is necessary in order to determine the specific relationships between distinct features of reward and punishment sensitivity, versus a cumulative index of reinforcement sensitivity, and ASD.

It remains unclear how individual differences in arousal or reward sensitivity mediates the relationship between BIS/BAS function and the different domains of ASD features. Here, we find several significant relationships between SPSRQ-C and SRS scores. Although the SRS is thought to be primarily a measure of social symptoms, it does contain an RBRI subscale. The RBRI subscale of the SRS was correlated with both SPSRQ-C Punishment and Reward; however, the relationship between SPSRQ-C Reward and the RBRI subscale was not found to be significant in the subsample (in Experiment 2). Thus, it’s unclear as to whether BIS/BAS function is related to a cumulative sum of ASD symptoms (i.e. overall symptom severity), specific symptom domains (i.e. social features or repetitive behaviors), or another complex behavioral phenotype that goes beyond traditional diagnostic categories (i.e. ASD versus non-ASD). Previous work in ASD has suggested that BIS/BAS function is primarily related to social features (Chevallier et al., 2012; Kasari et al., 1990; Mundy et al., 2007; Mundy, 1995) but not necessarily the features of RBRI. However, assessments of BIS activity in the context of ADHD/ODD symptoms have been linked to non-social behaviors (Geurts et al., 2009; Luman et al., 2012; Yerys et al., 2009). Future studies should continue to dimensionally assess sensitivities to punishment and reward in order to better understand which aspect of the BIS/BAS may differentially scale with pupil metrics and the range of social and non-social impairments seen in ASD.

While the current research highlights a linear relationship between features of reward sensitivity, ASD traits, and PD, a curvilinear or other nonlinear functional form should be explored as an alternative interpretation of the relationship between BIS/BAS activity and ASD features. Adapted versions of the classic Yerkes-Dodson law, describing the inverted U-shape relationship between arousal and behavioral performance, have been proposed to describe the neurocognitive development of reward processing regions (Van Leijenhorst et al., 2010) and the function of reward circuitry associated with clinically significant behavioral traits in ADHD (Plichta & Scheres, 2014). In the current research, our limited sample size left us underpowered to explore a multivariate, nonlinear model of arousal via PD, reward sensitivity, and ASD symptomology. Additional research in much larger samples is necessary in order to comprehensively quantify (a) the functional form of the relationship between reward sensitivity and core symptoms of ASD and (b) how this relationship is reflected in pupil measurements.

There are additional limitations in the present study that should be acknowledged and addressed as a part of future research. A large proportion of the individuals in our sample with an ASD diagnosis were male. Sex differences in ASD features and punishment and reward sensitivity were not central to the current research; however, sex differences in core clinical features, comorbid symptoms and in phenotypic variability continue to be a relevant topic across research in ASD. Additional research in much larger samples with representative proportions of both sexes is necessary in order to comprehensively characterize the manner in which motivational systems may be differentially related to autism features in males and females. Our sample also included a wide age range. Pupil size is linked arousal state, which can be impacted by age and other demographic variables (Loewenfeld, 1999). Additional research is needed to assess age-related effects on pupil dynamics and behavioral features of reward response associated with ASD. We also chose to use the SPSRQ-C, but other measures of reward sensitivity exist (Carver & White, 1994; Van den Berg et al., 2010). Future work should assess whether these results are consistent across different measures of reward sensitivity.

The current research makes a substantial contribution to current knowledge regarding punishment and reward sensitivity and ASD. Our results underscore the significant contributions of behavioral features outside of the core diagnostic criteria of ASD to the development and expression of clinically relevant features. This work also emphasizes that eye tracking technology can capture peripheral measures in patients with below and above average cognitive ability (Russell et al., 2019), indicating promise for future use in clinical trials of heterogeneous populations. Quantitative approaches are necessary in order to comprehensively characterize meaningful variability across behavioral features in order to identify factors that may moderate clinical outcomes across the ASD phenotype.

## Data Availability

That data described in the manuscript are not currently openly available.

## AKNOWLEDGEMENTS

The authors are grateful to Kayleigh M. Adamson for their help with recruitment and data collection. This study was funded by the Simons Foundation, SFARI Explorer Award #350225, as well as a Research Accelerator Award from the Autism Science Foundation.

## ETHICS APPROVAL & INFORMED CONSENT

All procedures performed in studies involving human participants were in accordance with the ethical standards of the institutional and/or national research committee and with the 1964 Helsinki declaration and its later amendments or comparable ethical standards. Informed consent was obtained from all individual participants included in the study. Specifically, parents and/or caregivers of participants provided informed consent for their child’s participation in the current study. Parents and/or caregivers also provided informed consent for their own participation as parents were asked to complete parent-report behavioral questionnaires as a part of the study design. When possible, assent was also provided directly from child participants using assent forms approved by the home insitution’s Insitutional Review Board (IRB). subset of individuals with ASD Dx (i.e. n=43 of N=89).

## Footnotes

1 Despite the fact that SPSRQ-C scores did not deviate from a normal distribution, we also evaluated the distribution of SPSRQ-C scores within our sample. SPSRQ-C Punishment scores appeared unimodal and were only mildly skewed, 0.351 (SE= 0.255). SPSRQ-C Reward scores also appeared unimodal with no indication of skewness, −0.095 (SE= 0.255). We assessed group differences in SPSPRQ-C scores in individuals with an ASD diagnosis versus those without. SPSRQ-C Punishment scores were significantly higher in those Individuals with an ASD diagnosis as compared to those who did not have an ASD diagnosis, t(87)=-2.33, *p*=0.022. SPSRQ-C Reward scores did not differ between individuals with and without an ASD diagnosis, t(87)= −1.31, *p*=0.192, NS.

2 To be included in analyses, we required participants to maintain fixation on screen during the baseline period and successfully complete at least 50% of trials across dark and light conditions in our eye tracking task. After excluding 16 participants, the average percent of trials successfully completed was 76.7 ± 15.1 (average with ASD diagnosis = 72.7 ± 15.6; average without ASD = 82.6 +/− 12.4; t(71)= −2.84, *p*=0.002). Unusable trials were due to failure of tracking caused by child noncompliance (closing eyes or averting gaze away from screen) or recording failure during the task.

